# Genomic and epigenomic evolution of metastatic prostate cancer: the first warm autopsy in China

**DOI:** 10.1101/2023.08.22.23294351

**Authors:** Wenhui Zhang, Yan Wang, Min Qu, Haoqing Shi, Xin Lu, Qingsong Yang, Fang Liu, Tao Wang, Ziwei Wang, Bijun Lian, Ling Chen, Xiaoyi Yin, Yongwei Yu, Jing Li, Xu Gao, Zhuan Liao

**Affiliations:** Department of Urology, Changhai Hospital, Second Military Medical University, Shanghai 200433, China; Department of Radiology, Changhai Hospital, Second Military Medical University, Shanghai 200433, China; Department of Urology, the First Affiliated Hospital of Zhengzhou University, Zhengzhou, Henan 450052, China; Department of Urology, the 903rd PLA Hospital, Hangzhou, Zhejiang 310012, China; Department of Oncology, Changhai Hospital, Second Military Medical University, Shanghai 200433, China; Department of Hepatobiliary Pancreatic Surgery, Changhai Hospital, Second Military Medical University, Shanghai 200433, China; Department of Pathology, Changhai Hospital, Second Military Medical University, Shanghai 200433, China; Shanghai Key Laboratory of Cell Engineering, Second Military Medical University, Shanghai 200433, China; Department of Bioinformatics, Center for Translational Medicine, Second Military Medical University, Shanghai 200433, China; Clinical Research Center, Changhai Hospital, Second Military Medical University, Shanghai 200433, China

**Keywords:** Clonal evolution, Intratumor heterogeneity, Prostate cancer, Metastasis, Epigenetics

## Abstract

**Background:** The development and expansion of warm autopsy program have important implications in dissecting the heterogeneity during cancer dissemination and resistance. However, in China, the practice of warm autopsy has not yet been officially launched and documented.

**Methods:** To explore and establish the procedures and standards for warm autopsy in China, we followed the disease course of a male patient with terminal metastatic prostate cancer. We assembled a multidisciplinary team to perform warm autopsy immediately after death. Through longitudinal sampling from biopsy and autopsy, we performed integrative and comprehensive genomic and epigenomic analysis using multi-omics approaches.

**Results:** We traced the dynamic evolution and heterogeneity of this prostate tumor, and identified many critical driver events in both the original tumor and its disseminations. Truncated *CDKN1B* may result in downregulation of expression, which represent a key driver event in the metastatic progression of prostate cancer. We also delineated the congruence of genetic and epigenetic clonal evolution during tumor metastasis.

**Conclusions:** Our data and analysis elucidated the mechanisms and drivers during metastasis, which represent a valuable resource for the study and treatment of prostate cancer. We also call on more investigators to improve warm autopsy of prostate cancer for clinical and experimental investigations.

## Background

Prostate cancer is the one of the most common malignancies of men in China, and the incidence and mortality rates are much higher than the average levels worldwide^1^. The majority of prostate cancer deaths are attributed to metastatic dissemination of the primary tumor^2,3^. The collection and investigation of tumor samples at multiple metastatic sites can be instrumental in dissecting the evolving complexity of tumor metastasis. The needle biopsy is not practical for simultaneously acquiring metastatic tumors from multiple sites of the same patient. In contrast, multiple tissue harvesting by autopsy is an excellent way to collect most tissues and preserve most viable samples.

The Autopsy Section investigates final diagnosis of the disease which caused the death of a patient in clinic. In academic settings, autopsies provide an opportunity for researchers and students to understand disease process by studying postmortem tissues and the clinical record. An important function of an academic autopsy service is to process tissues for research purposes. The advent of “immediate” autopsy in 1976 enabled acquisition of inaccessible tissues and cells for culturing^4^. The prototype of “warm autopsy” at an academic institution was developed in the 1980s^5^. After the stable and integrated infrastructure setting up for cells isolation, the purpose for rapid autopsy had been shifted to specimen biobanking^6^. In 2000, “warm autopsy” as a term has been defined as acquisition of tissue samples through immediate autopsy shortly after death^7^. A team at University of Michigan successfully collected 8 different tissue types from 14 warm autopsy cases within 4 years, meeting the need of collecting high-quality primary and metastatic tumor samples for molecular studies.

Today, with the advent of next-generation sequencing, warm autopsy represents the next phase in modern pathology. Warm autopsy program has now been implemented in many countries^8–10^. However, the study of warm autopsy has lagged behind in China. There is no documented program for this specific research area in China to-date.

Here, we report the first, to our knowledge, warm autopsy in China. By performing sequencing on eight metastatic tissues collected via warm autopsy from an advanced prostate cancer patient, we revealed extensive intratumor heterogeneity between different metastases, and delineated the congruence of genetic and epigenetic clonal evolution. Our work developed the procedures and standards for warm autopsy, and we call on Chinese urologists and oncologists to promote warm autopsy of prostate cancer for clinical and experimental investigations.

## Methods

### Patient information and course of the disease

A male patient in his 60s was diagnosed with prostate adenocarcinoma in January 2016. The transrectal ultrasound-guided prostate biopsy showed prostate cancer (T3aN1Mx, tPSA: 20.166 ng/ml, Gleason Score: 5+5). The patient was started on treatment with ADT and chemotherapy, after a short partial response (PSA 2.52ng/ml). Soon after the patient got into mCRPC. Left clavicle lymph node and multiple vertebral metastases were detected by MRI in July 2016. Then the ADT and chemotherapy were suspended, and collection of plasma and lymph node (LN) biopsy was initiated for further treatment evaluation. Two months later, treatment was switched to Vemurafenib due to a *BRAF* V600E mutation detected by the ctDNA targeted sequencing. General deterioration then occurred, with disease progression into cachexia (dyspnea, renal insufficiency and heart failure). Treatment was stopped and the patient died on January 2017. Tumor samples for sequencing were obtained from the primary prostate sites at diagnosis and metastases at warm autopsy.

### Committee establishment

With the support of the Changhai Warm Autopsy Team (CWAT) and Red Cross, the committee of warm autopsy was established, which consisted of a clinical care team (including urologists and oncologists), surgeons (including general surgeons, orthopedists, cardiothoracic surgeons, and neurosurgeons), pathologists, researchers (including translational medicine center and laboratory assistants), ethics board and the administrative unit (Fig 1). Patients diagnosed with end-stage CRPC were referred to the Urology Department of Shanghai Changhai Hospital. The objectives and procedures for this study were explained to the patient carefully. Permission for warm autopsy was obtained before the death, with consent provided by the patient and the family members. After death, consent was obtained by the family members with the witness by the operator and the ethics committee. Written informed consent was obtained in accordance with Chinese legislation. Sample collection and study were approved by the ethical board at Changhai Hospital (No: TMEC2014-001, No: CHEC2019-165).

**Fig. 1.**
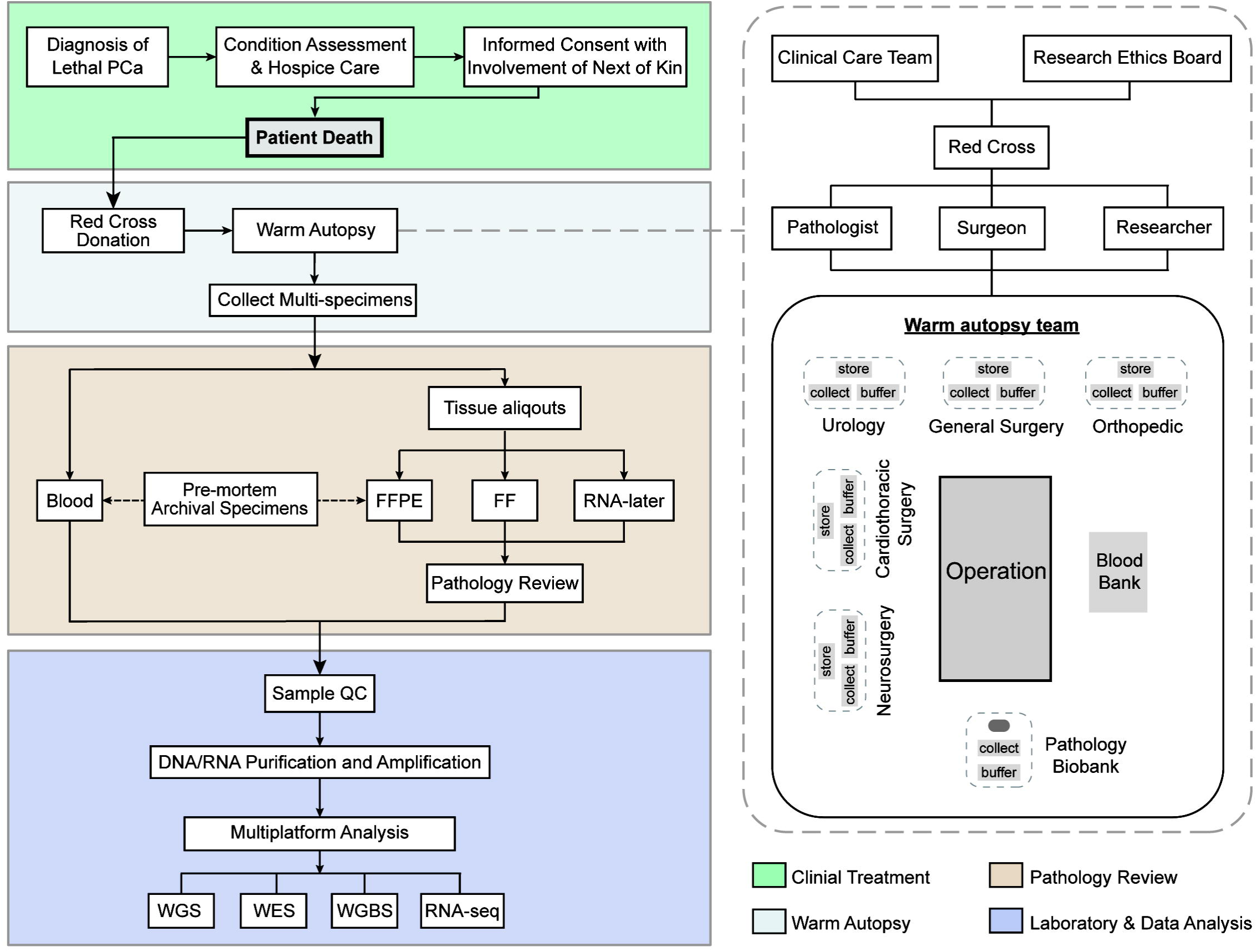
Workflow diagram giving overview of warm autopsy. The four steps of the study are shown on the left. The right shows in detail the organization of the warm autopsy committee, as well as the operating table layout in implementation.

### Implementation of the warm autopsy

The body was sprayed with ethanol before operation. Then the operator approached the internal organs through an “+ shaped” incision, up to the collarbone, down to the pelvis, extend the middle route on both sides. The skin and subcutaneous tissues were retracted from the thorax and abdomen, and the blood and other body fluid were collected in the tubes. Around the warm autopsy operator, there were 6 independent collection areas for tissue procurement and storage (Neurosurgery, Cardiothoracic, Urology, General Surgery, Orthopedic, and Pathology biobank). Firstly, the urological operator resected the prostate, urinary bladder, and pelvic lymph nodes in a manner similar to that of a cystoprostatectomy. Then the kidneys, adrenal glands and ureters were resected in a manner similar to that of a nephroureterectomy. Secondly, the breastplate was removed by cardiothoracic surgeon. The heart, lungs, aorta and mediastinum were resected orderly. Meanwhile, the general surgeon reseed the stomach, liver, gallbladder, pancreas, colon and retroperitoneal lymph nodes. Thirdly, the orthopedist collected the bone metastases as well as the control tissues, and the neurosurgeon collected the brain. The tissue was cut into approximately 10mm×10mm×20mm slice, which was divided into three contiguous aliquots for the homology of purified DNA/RNA/protein. And we do triplication repeat to guarantee the quality of tissue. Finally, tumor tissues from eight metastatic sites and normal tissues from 23 anatomic sites were procured by warm autopsy. The collection of normal tissues referenced the Genotype-Tissue Expression (GTEx) project^11^, included kidney (cortex), ureter, bladder, testis, adrenal gland, brain (cortex), brain (cerebellum), bone, heart (left ventricle), artery, breast, thyroid, esophagus (muscularis), stomach, pancreas, omentum, gallbladder, liver, colon, ileum, jejunum, muscle, skin (suprapubic).

### Pathological evaluation of specimens and tissue storage

All autopsy samples were immediately frozen in liquid nitrogen and stored at −80 °C. One primary sample (PB2) and seven fresh-frozen samples from warm autopsy except for bone (LV2M) were also stored in RNAlater (Ambion) for RNA extraction according to manufacturer protocol (Supplementary Table S1). Hematoxylin and eosin (H&E) stains were performed for thin slices of snap-frozen, OCT-embedded tissue blocks using standard techniques. Tumor staging and pathological grading were independently determined by two professional uropathologists. Bone metastases were defined using emission computed tomography (ECT). According to EAU guidelines, the diagnostic criteria for CRPC were the castrate levels of serum testosterone < 50 ng/dL, or 1.7 nmol/L plus one of the biochemical progression or radiological progression^12^.

### Somatic mutations calling

Somatic mutations including single nucleotide variants (SNVs) and small-scale insertions/deletions (Indels) were detected using Mutect2 from GATK v4.0.11.0 and Strelka2 v2.8.2^13^. The annotation of the somatic mutations was performed with the ANNOVAR tool ^14^. In addition, we used the panel of normals (PoNs) built from the 208 normal samples in the previous study^15^ to remove and filter the recurrent technical artifacts that mutations appeared in more than one PoN samples. We removed mutations with minor allele frequency > 5% in the NHLBI GO Exome Sequencing Project (http://esp.gs.washington.edu), the 1000 Genomes cohort (http://www.internationalgenome.org), and the ExAC resource (http://exac.broadinstitute.org), but retained all COSMIC variants. Mutations within the blacklist that we compiled according to the MutSigCV paper were also filtered and removed^16^. Additionally, we also removed variants in repetitive elements (RepeatMasker http://www.repeatmasker.org/) or segmental duplications (UCSC hg19.genomicSuperDups). Finally, the candidate mutations in previously implicated cancer genes were reviewed manually on the Integrated Genomics Viewer (IGV) to ensure that no candidate driver mutations were mistakenly removed. The correct nomenclature for mutation was checked applying Mutalyzer^17^.

### Somatic copy number alterations calling

We utilized the FACETS v0.5.6 algorithm^18^ to detect somatic allele-specific copy number alterations (CNA) and determine the tumor purity of the tumor samples from WGS data. The copy number alteration events were defined to meet the following criteria as previously described^19^: (1) regions with total copy number = 3-8 were defined as a gain event; (2) total copy number > 8 were defined as an amplification event; (3) regions with total copy number = 1 were defined as a loss event, and (4) total copy number = 0 were defined as a homozygous deletion event. Additionally, regions with minor copy number = 0 and total copy number > 0 will be defined as a loss of heterozygosity (LOH) event, including LOH (or loss, total copy number = 1), CN-LOH (Copy neutral loss of heterozygosity, total copy number = 2) and Gain-LOH (Copy gain loss of heterozygosity, total copy number > 2).

### Genetic clone evolution analysis and driver events annotating

PyClone^20^, an algorithm to infer clonality by the Bayesian clustering method, was conducted to reconstruct the accurate clonal and subclonal architecture across primary and metastatic tumors. The somatic mutations with depth higher than 30× for WGS and 100X for WES were used as input to PyClone. Using the Bam-readcount v0.8.0 (https://github.com/genome/bam-readcount) with minimum base quality 20, we calculated the reference and alternate read-depth of each mutation from the BAM files. To infer cancer cell fraction (CCF) and cluster mutations, PyClone beta-binomial model with the “parental_copy_number” and “--tumor_contents” option was run for 50,000 iterations. Clusters containing two or fewer variants were discarded. Mutations clusters identified were imported into ClonEvol ^21^ to track and visualize tumor’s clonal evolution (Supplementary Table S2). The potential driver events for somatic mutations and CNA were annotated as described previously^19^. Driver gene mutations were labeled in clonal evolution trees according to the results of ClonEvol, and the driver CNA events were labeled by matching the presence/absence status of the copy number events with that of the clusters across samples. The body map was drawn by referring the study of Gunes Gundem et al^22^.

### Analysis for Intratumor heterogeneity of DNA methylation

Methylation calls and differentially methylated cytosines were analyzed using R package methylKit v1.2.0^23^. No batch effects were identified by assocComp in methylKit and there were no plating issues. The top 1% of CpG sites (n=150,000) with the greatest intratumoral methylation range were selected for downstream analysis. Clustered heatmaps were drawn using the superheat package (https://github.com/rlbarter/superheat), and Euclidean distance was calculated for hierarchical clustering. We calculated the DNA methylation variability^24^, which was defined by the median of the range of CpG sites, to measure the intratumoral heterogeneity of methylation in genomic contexts. The higher the methylation variability represents the more ITH observed. The CpG annotation reference was obtained from the R package annotatr (https://github.com/hhabra/annotatr). Methylation levels at least 20% increase/decrease relative to the average of normal samples were defined as hyper-methylation and hypo-methylation, respectively.

### Reconstruction of methylation and mutation-derived phylogenetic tree

We performed the dist function in the R to generate DNA methylation Euclidean distance matrices using the top 1% of CpG sites with the greatest intratumoral methylation range. The mutation Euclidean distance matrices were calculated using the CCF or VAF of mutations that were input to ClonEvol, and the mutations private to PB2 and LV2M were removed. The fastme.bal function in the R package ape^25^ was used to reconstruct the Euclidean distance matrices-based phylogenetic tree by applying the minimal evolution algorithm. Confidence for the branches on phylogenetic trees was assessed by bootstrapping (1,000 bootstrap replicates) using the boot.phylo function in the R package ape. Congruence between the genetic and epigenetic distance matrices was calculated by the Spearman’s correlation coefficient.

More detailed methods are provided in Supplementary Materials and Methods.

## Results

### Program development: the warm autopsy in China

To gain insight into the intratumor heterogeneity and evolutionary mechanisms during tumor metastasis and drug resistance, we established the Changhai Warm Autopsy Team (CWAT), a multidisciplinary team, to recruit cancer patients close to the end of life and perform warm autopsies for tumor sampling in disseminated disease (Fig 1). The team was consisted of urologists, general surgeons, orthopedists, cardiothoracic surgeons and neurosurgeons for harvesting all normal and cancer tissues; blood bank technicians for blood collection; additional members responsible for tissue processing and labeling; pathologists for pathology review and translation medicine center personnel for research and data analysis. All team members were permanent employees and were available round-the-clock. The warm autopsy committee approved the protocol, procedures, and methods for obtaining informed consent. In Fig 1 we diagramed the warm autopsy operation station, which contained 6 independent collection areas for the tissue procurement and storage. The unique identifier of each sample was assigned during autopsy. All clinicopathologic information related to the patient’s medical and surgical history, medications, imaging studies, pathology and autopsy findings were deposited in a follow-up database PC-Follow^TM^ ^26^.

### Patient information and tissue collection

The subject was diagnosed with prostate adenocarcinoma, and received androgen deprivation therapy (ADT) combined with systemic chemotherapy right after primary diagnosis (Fig 2A). However, an elevated prostate specific antigen (PSA) level and multiple metastases detected by imaging indicated disease progression to metastatic castration-resistant prostate cancer (mCRPC) in July 2016. To assess tumor progression, ADT combining chemotherapy was discontinued, and the metastasis was found at left supraclavicular and right pelvic lymph nodes by ultrasound-guided needle biopsy. Targeted sequencing was performed in plasma samples using a custom gene panel for ctDNA (circulating tumor DNA), and Vemurafenib therapy was applied based on the discovery of the *BRAF* V600E mutation. The allele fraction (AF) of *BRAF* mutation decreased during targeted therapy (Supplementary Fig 1). After a slight improvement in the patient’s condition, dyspnea and renal insufficiency began to appear in December 2016. Despite systematic support, the patient succumbed to multiple organ failure in January 2017. After death, the tumor tissues from eight metastatic sites and normal tissues from 23 anatomic sites were procured and stored by warm autopsy (Fig 2B). The autopsy was completed in a total of 2.5 hours, and performed as outlined in “Methods”.

**Fig. 2.**
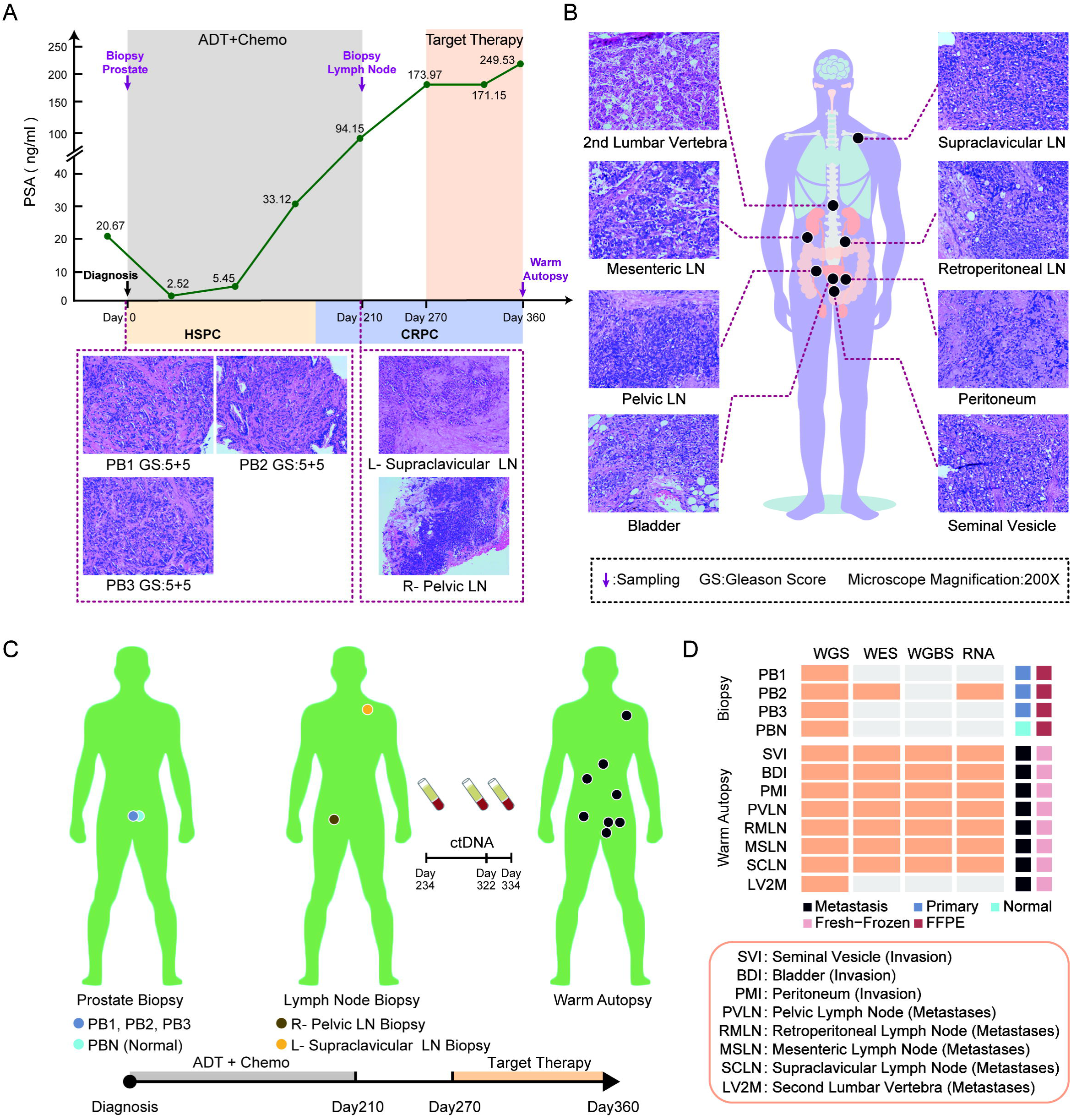
Clinical course and serial procurement of the patient. (A) The clinical course of disease progression and treatment in this index case. The bottom figure shows the pathology of biopsies (200x magnification). (B) The pathology of metastatic sites procured by warm autopsy (200x magnification). All the hematoxylin and eosin (H&E) stains were performed using standard techniques. (C) Tissue and blood samples taken at different times. Two lymph node metastases samples taken on day 210 were not sequenced. R, right; L, left; LN, lymph node. (D) The type of sequencing performed. The abbreviations of samples are indicated. FF, fresh frozen; FFPE, formalin-fixed paraffin-embedding.

In this patient, the autopsy revealed widely disseminated prostate tumors in bladder, peritoneum, seminal vesicles, lymph nodes and bones (Fig 2B, Supplementary Fig 2A). Among them, we observed that the chains of lymph nodes were involved by metastatic spread, and the retroperitoneal and supraclavicular lymph nodes were two obvious sites of metastasis. Additionally, we identified additional metastases through autopsy and pathological review; however, due to the limited sample size and ethical considerations, we did not retain them for further analysis (Supplementary Fig 2B). In addition to common metastatic sites of prostate cancer such as the adrenal gland and liver^27,28^, we also identified the metastatic cancer thrombus in quadriceps femoris muscle and inferior vena cava, and skin metastases (Supplementary Fig 2B). These rare and atypical metastatic foci highlight the complexity of prostate cancer dissemination.

To determine the genetic and epigenetic heterogeneity and metastatic progression of cancer that gave rise to the lethal tumor burden, high quality tumor samples collected across different clinical courses were profiled using multi-omics approaches including whole genome sequencing (WGS), whole-exome sequencing (WES), whole-genome bisulfite sequencing (WGBS) and RNA sequencing (RNA-seq) (Fig 2C, D). In total, 11 tumor samples and one non-cancerous matched healthy prostate tissue were collected for sequencing. The tumor samples included three treatment naïve tumor specimens were obtained from prostate FFPE biopsy, and eight metastases from warm autopsy (Fig 2D). To describe the heterogeneity of primary tumor, we performed WGS on four prostate samples. Limited by the small size of the puncture tissue, however, we only selected an adequate sample (PB2) for additional WES and RNA-seq. For seven metastases samples, we performed WGS, WES, WGBS and RNA-seq to dissect the evolutionary progression, while bone samples (LV2M) only performed WGS due to the small sample size (Supplementary Table S1).

### Intratumoral genetic heterogeneity of prostate primary tumors

To characterize the genetic origin and subclonal architecture of primary tumors, we analyzed WGS data of three biopsy samples at the time of initial diagnosis (Fig 2C). Due to the relatively low tumor purity (34%) of sample PB2 estimated by WGS, we further performed ultra-deep WES (1,119X) on this sample. All 3 treatment-naïve tumors shared widespread somatic mutations, including *TP53* p.(Asn179Glnfs*26) and *CDK12* p.(Ala993Val) (Fig 3A) that have been demonstrated to be frequently mutated in previously characterized metastatic prostate cancer cohorts^29,30^. To determine the clonality of somatic mutations, we further calculated the AF distributions (Fig 3B). Remarkably, the *TP53* and *CDK12* mutations had the largest allele frequencies which matched each tumor sample’s purity, suggesting that these mutations are clonal and present in almost all tumor cells. Copy number alteration (CNA) analysis revealed that all three samples underwent loss of heterozygosity (LOH) of *TP53* and *CDK12*, resulting in complete inactivation of these known prostate cancer genes (Fig 3C). Utilizing the somatic mutations as a molecular clock as described previously^31^, we explored the sequential accumulation of LOH events of these two driver mutations (Fig 3D). For example, the *TP53* mutation clearly occurred before the 17p copy neutral loss of heterozygosity (CN-LOH), as it was present on both copies of chromosome 17p. Although it’s hard to infer the exact order of *CDK12* mutation and one copy loss of chromosome 17q11.2-21.2 by existing algorithms, the conclusion of *CDK12* and *TP53* mutation occurred before tumor clonal expansions could be drawn.

**Fig. 3.**
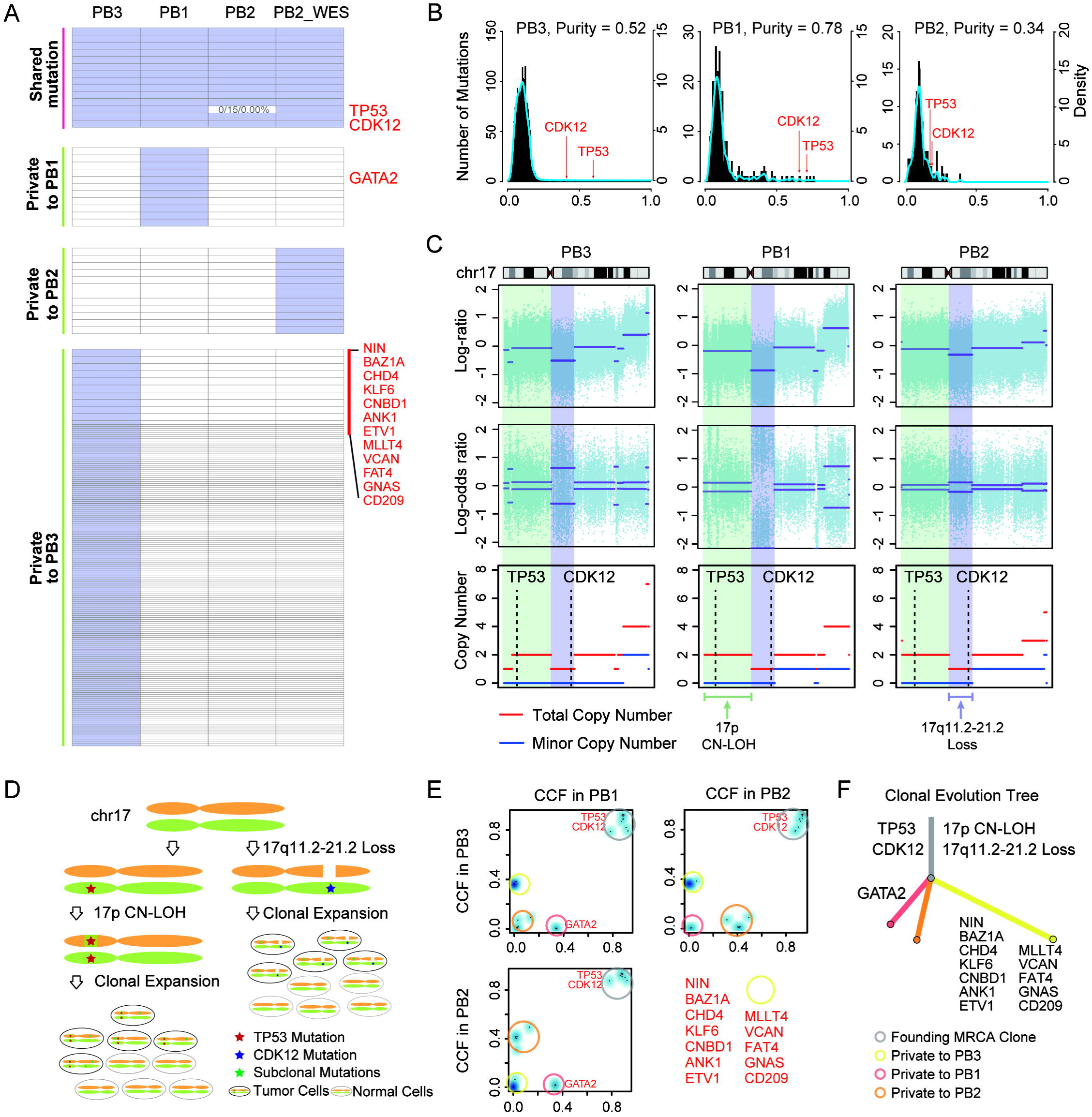
Intratumoral genetic heterogeneity of prostate primary tumors. (A) The regional distribution of nonsynonymous mutations in primary tumors. The heat map indicates the presence of a mutation (purple) or its absence (white) in the individual tumor. Right showed the gene names of driver mutations. The *TP53* mutation was not detected in the PB2 sample using WGS due to insufficient total depth, and the number in square indicate the alt_reads/total_reads/allele_fraction. (B) The figure shows the allele fraction distributions plotted by mutation number (left vertical axis) and density (right vertical axis). Tumor purity provided by FACETS in three samples is also indicated. (C) Copy number profile of chr17 and the LOH of *TP53* and *CDK12*. Shown from upper to lower are the total copy number log-ratio (the log ratio of total read depth in the tumor versus that in the normal), allele-specific log-odds-ratio (the log odds ratio of the variant allele count in the tumor versus in the normal), and corresponding integer (total, minor) copy number calls provided by FACETS. (D) The figure demonstrates how somatic mutations accumulate in a CN-LOH (*TP53*) and Loss (*CDK12*) chromosome. (E) Cancer cell fractions and clusters of mutations inferred by PyClone for pairs of samples. Blue density areas reveal the mutation clusters present at clonal or subclonal levels, and the manually colored circles provide the localization of mutation clusters in different samples. Driver mutation genes present in the cluster are marked in red. (F) The clonal evolution tree of the primary tumor. The length of branches connecting clones is proportional to the number of mutations contained, and the driver events identified are marked on the tree.

Next we determined the clonal relationship between tumor cell subpopulations. We calculated the cancer cell fractions (CCF) of mutations between pairs of samples, and constructed the clonal evolution tree based on somatic mutations using the previously published methods (Fig 3E, F). We identified subclonal mutations private to each of the three primary tumors, all were seeded directly from the most recent common ancestor (MRCA) harboring the *TP53* and *CDK12* alterations. If we used the number of mutations in the corresponding subclones to represent branch lengths in the evolution tree, we found that subclone of the PB3 tumor acquired significantly more mutations than PB1 and PB2 (Fig 3A, F). However, examining the mutational signature of PB3 did not reveal meaningful interpretations for this anomaly (Supplementary Fig 3). Manual review of these mutations confirmed that they are true positive mutations for PB3, and not false negatives for PB1 or PB2. This result highlights the intratumoral heterogeneity of primary prostate tumor. Overall, we observed inactivation events of *TP53* and *CDK12* genes in early tumor evolution stage, suggesting that they are driver events for the origin of prostate cancer in this patient.

### Intratumoral genetic heterogeneity of prostate metastatic tumors

Warm autopsy could provide an invaluable resource to determine the mutational landscape and cancer evolution from localized tumor to metastatic tumors, and to reveal potential treatment targets. To understand the genomic intratumoral heterogeneity, we applied stringent alterations calling pipelines including somatic mutations and copy number alterations (CNAs) using WGS data from all sequenced samples (Fig 2D). In total, an average of 96 (25–238) and 57 (41–120) nonsynonymous mutations were detected in primary and metastatic tumors, respectively (Fig 4A). At the CNA level, an average of 98 (31–213) gains or amplifications, 38 (8–68) losses or deletions and 23 (8–62) CN-LOHs were detected. The average tumor purities of all samples were 0.60 (0.34-0.78) as estimated by FACETS^18^. We defined the driver events (Fig 4B, Supplementary Table S3) for each tumor using previously established methods^19^. In addition to the clonal mutations in *CDK12* and *TP53*, amplification of the AR gene was observed in all metastatic samples which could contribute to castration resistance (Fig 4B). Intriguingly, a truncating mutation of *CDKN1B* p.(Gly97Valfs*22) private to all metastases except for bone sample (LV2M) was identified and validated (Supplementary Fig 4). These findings suggested that the truncated CDKN1B may be a potential driver for tumor lymphatic metastases and periprostatic organ invasion. We also conducted preliminary data analysis and experimental verification in the subsequent section.

**Fig 4.**
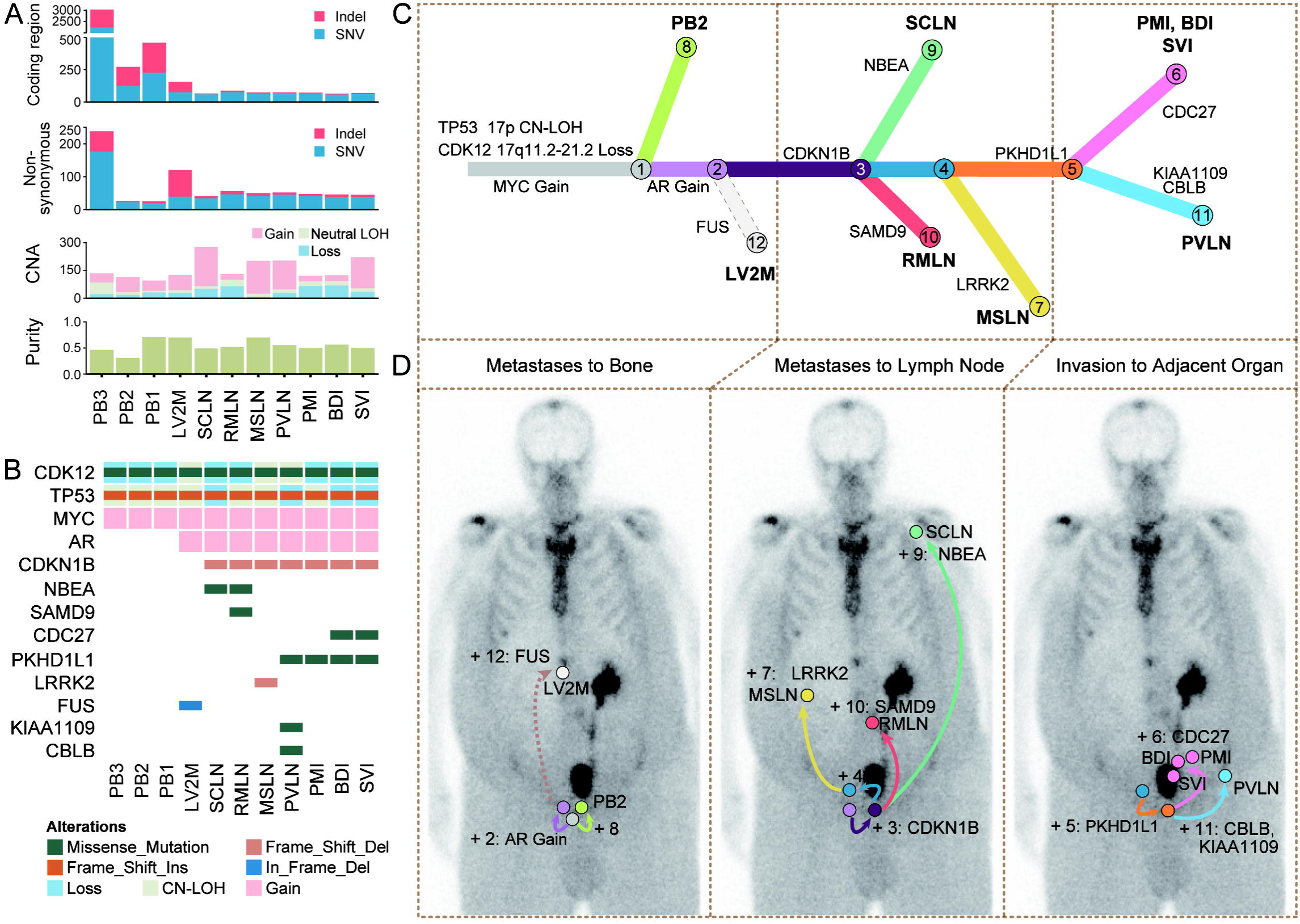
Intratumoral genetic heterogeneity and clonal evolution of prostate metastatic tumors. (A) An overview of somatic alterations detected in 11 tumors. Each panel displays the number of mutations in coding region, nonsynonymous mutations, the number of segments for copy number alterations, and the tumor purity, respectively. (B) Overview of the analyzed driver genomic alterations in the primary tumor and metastases. (C) The clonal evolution tree of the primary tumor and metastases inferred by ClonEvol. Except for cluster 12 private to LV2M, which is manually added, all the CCF clusters were calculated by PyClone. The branch length is scaled by the log2 ratio of the number of mutations in the individual clone. The potential driver events are highlighted. (D) The emergence and movement of clones in the spread of metastasis. The color-coded arrows depict the seeding events and the acquisition of mutations, and the sequence of events is ordered according to the clonal evolution relationship. Plus (+), the acquisition of subclone.

To infer the clonal architecture in tumor metastasis, we utilized mutation and CNA information from WES data which had higher depth to perform clonal evolution analysis. We selected 285 somatic mutations detected by both WGS and WES (except for LV2M) as input to PyClone^20^ and identified 11 major clusters by calculating the CCF and clustering (Supplementary Fig 5). The clonal ordering and a phylogenetic tree were constructed by ClonEvol^21^, which determined the clonal relationship among tumors (Fig 4C). We observed the tumor metastasis underwent approximately three processes that are distinctly histologically characterized. The tumor was first metastasized to the bone (LV2M), and then spread to the lymph nodes (SCLN, RMLN, and MSLN) in turn after acquiring the metastatic dominant mutation of *CDKN1B*, and finally invaded the adjacent organs of the prostate (PMI, BDI, and SVI). The “body maps” were created to characterize the order of driver events and movement of subclones from primary tumor to metastasis (Fig 4D). Metastasis to bone was directly seeded by MRCA, suggesting that LV2M may appear early in systemic disease with a distinct pathway from other metastases.

To summarize, from a genetic and molecular perspective, tumor metastases in this case was primarily driven by *CDKN1B* mutations, but bone metastasis was characterized by an entirely different evolutionary process harboring *FUS* mutation.

### *CDKN1B* alterations in prostate cancer metastasis

To delineate the genomic alteration atlas of *CDKN1B* in prostate cancer, we preliminarily checked alterations of *CDKN1B* in prostate cancer cohorts from Cbioprotal (https://www.cbioportal.org/). Among a total of 9510 prostate cancer patients, approximately 4% exhibited variations in the *CDKN1B* gene, which were predominantly truncated mutations and deep deletions (Supplementary Fig 6A). We checked the mutational spectrum, and found the *CDKN1B* mutation p.(Gly97Valfs*22) detected in this case were also identified by the metastatic prostate cancer cohort MSK^32^ (Fig 5A). Importantly, we observed the mutation was located within a hot spot (amino acids 83-102), which specifically enriched in the metastatic patients. Even though we did not obtain statistically significant results, these findings still indicated that hot spot mutations represented by p.(Gly97Valfs*22) may be driver events leading to prostate cancer metastasis. Furthermore, using RNA-seq data from this warm autopsy patient and previously published CPGEA cohort^15^, we found that the RNA expression of *CDKN1B* was down-regulated in metastases (truncating) compared with primary tumor samples (Supplementary Fig 6B). The pattern was further validated by using TCGA public data^33^ (Supplementary Fig 6C). These results implied that the genomic variations (truncating or deletion) of *CDKN1B* could lead to down-regulation of expression and promote the metastasis of prostate cancer.

**Fig 5.**
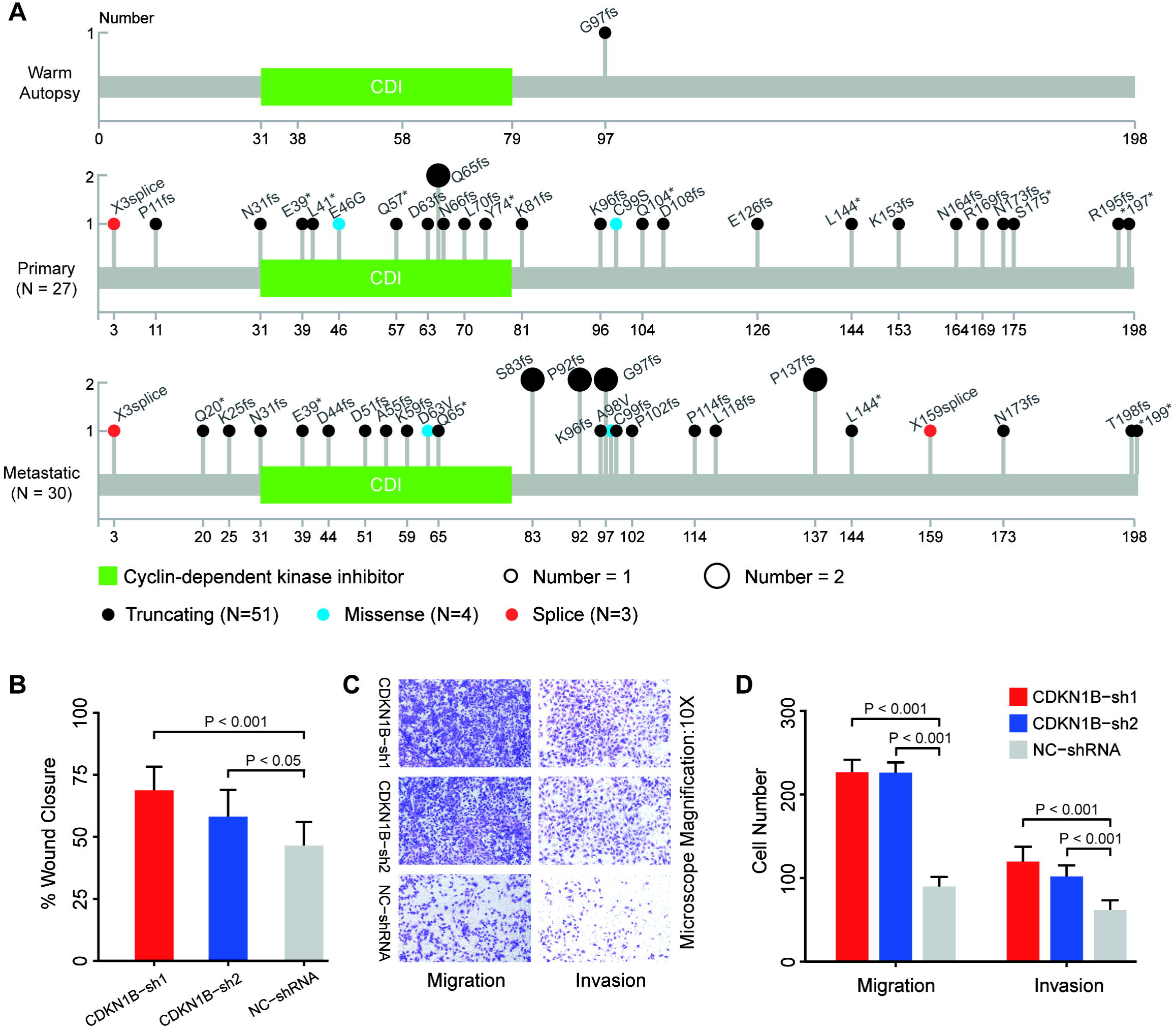
*CDKN1B* alterations in prostate cancer metastasis. (A) The position distribution of *CDKN1B* somatic mutations in this patient and in the deduplicated samples from cbioprotal. The circles are colored with respect to the different mutation types, and the size represents the number of patients with the mutation. (B) Scratch assay of 22RV1 cells transfected with shRNA (n = 3), which corresponds to Supplementary Fig 7D. (C, D) Images of migrating and invading cells tested by using Transwell assays for 22RV1 cells transfected with shRNA (n = 3). *P* values were determined by two-tailed Student’s t test.

To explore the function of the down-regulation of *CDKN1B* expression, we designed and synthesized two sets of shRNA sequences and one set of control shRNA sequences. The cell line 22RV1 was chosen in all subsequent experiments due to the highest expression of *CDKN1B* (Supplementary Fig 7) and originating from primary prostate cancer. We observed that the migration and invasion abilities in the *CDKN1B* downexpression group was significantly increased when compared with the control group (Fig 5B, C and D, Supplementary Fig 7). Taken together, the results suggested that the down-regulation of *CDKN1B* may be involved in metastasis of prostate cancer.

### Intratumor heterogeneity of DNA methylation and epigenomic evolution

Our previous study demonstrated the congruence of genomic and epigenomic tumor evolutionary histories in a large population^15^. To comprehensively dissect the epigenetic clonal evolution of tumor metastases, we applied genome-wide DNA methylation analysis from seven metastatic samples in this case (Fig 2D). The WGBS data of three normal prostate specimens from a previous study^34^ were used as the reference prostate epigenome in this analysis (Supplementary Fig 8A, B). By using the 1% of CpG sites with the greatest intratumoral methylation variation for unsupervised clustering analysis, we observed the overall hypomethylation patterns in tumor samples and the extensive epigenetic heterogeneity across anatomically distinct metastasis samples (Fig 6A). To define the signatures of DNA methylation heterogeneity, we examined the genomic context of the CpG sites with the most variable methylation levels across samples. The variable sites within tumors were not distributed evenly across the genome, but were enriched in CpG islands and promoters, which showed greater intratumoral heterogeneity than other genomic features examined (Fig 6B).

**Fig 6.**
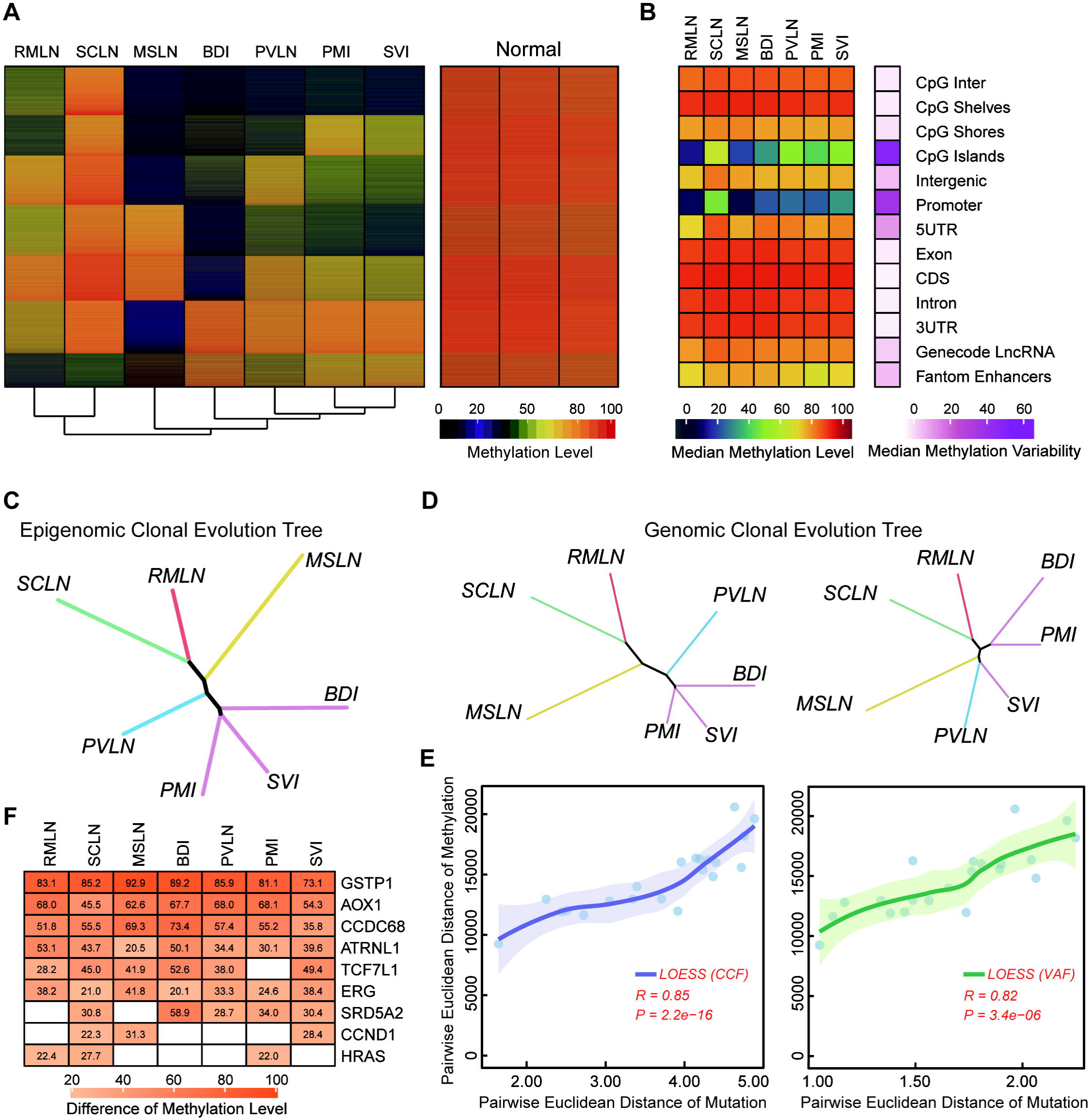
Intratumor heterogeneity of DNA methylation and epigenomic evolution in prostate metastatic tumors. (A) The figure shows the intratumoral heterogeneity of methylation patterns depicted by unsupervised hierarchical clustering using the top 1% of CpG sites (n = 150,000) with the greatest difference. Normal samples were excluded for the hierarchical clustering but were input into row clustering. (B) DNA methylation Intratumor heterogeneity on genomic regions. The median methylation variability on the right of the figure was calculated by the range of CpG sites (maximum level - minimal level) between tumors. (C) Epigenomic clonal evolution tree inferred from DNA methylation distance matrices. Lengths of trunks and branches were inferred using the top 1% of CpG sites (same as Fig 5A, see Supplemental Methods). Color coding is the same as in Fig 4F. (D) Genomic clonal evolution tree inferred from CCF (cancer cell fraction, left) and VAF (variant allele fraction, right) distance matrices. (E) The correlation between epigenomic distance matrices and genomic distance matrices (CCF, left; VAF, right). LOESS fitted curve and 95% confident interval are presented. Rho coefficient (*R*) and P value (*P*) are assessed by Spearman’s rank correlation. (F) The difference in methylation levels of CpG island in promoter region of known prostate cancer driver genes (www.genome.jp/pathway/ko05215) between each tumor and three normal prostate samples. Hyper-methylation and hypo-methylation were defined as difference of more than 20%. White cells in the heatmap represent differences below 20%.

The highly dynamic genetic alterations and the extensive epigenetic heterogeneity across metastases prompted us to investigate the association between genetics and epigenetics during tumor evolution. We constructed a phylogenetic tree based on DNA methylation levels across samples^24^. The epigenetic phylogenetic tree almost superimposed the genetic tree constructed based on genetic mutation data in their topology (Fig 6C, Fig 4C, Supplementary Fig 8D). To quantify the resemblance between somatic mutation and DNA methylation based phylogenetic trees, we used variant allele fraction and cancer cell fraction to calculate the Euclidean distance and constructed the genetic evolution trees, respectively (Fig 6D). In addition to CCF showing a phylogenetic relationship almost consistent with methylation, we also confirmed the congruence between epigenetic and genetic evolution by calculating the Spearman correlation coefficient for distance matrices (Fig 6E). Finally, we defined the hyper- and hypo-methylation of CpG island in promoter region of known prostate cancer driver genes, and investigated which genes were affected by methylation events (Fig 6F, Supplementary Fig 8C). We observed that some genes that have been verified to be frequently altered in DNA methylation in prostate cancer, such as *GSTP1*, *AOX1* and *CCDC68*^15,35,36^, were hypermethylated in all metastases in this patient, indicating that they were likely clonal events and may occur earlier in prostate cancer evolution. Together, we showed the extensive epigenetic heterogeneity across prostate cancer metastases, which displayed a phylogenetic relationship recapitulating the genetic clonal evolution.

## Discussion

The development of warm autopsy program empowered the investigators with the unprecedented ability to thoroughly sampling and comprehensively dissect the evolutionary trajectories of metastatic-lethal prostate cancer^7,22,37,38^. Warm autopsy program now has been implemented in various types of research facilities: non-profit research institutes, universities and federal research institutes in America^7,10^, Spain^39^, England^40^, Australia^41^, etc. However, protocols and guidelines for warm autopsy have been individually modified according to the policies of different countries, such as ethics^9^, standard operation procedures^8^ and difficulties in practice^5,10^.

Despite the potential benefits of warm autopsy, the barriers and restrictions in implementation and development have limited the number of autopsies in cancer patients, especially in China^42^. Due to the traditional concept of preserving the intact body after death, the development of the body donation and autopsy system in China has been relatively slow ^43^. The societal and community resistance are generally derived from concerns regarding body disfigurement after the autopsy, unwillingness from the patient’s families, and a lack of awareness towards the practice of autopsy. The logistical challenges not only require researchers to build a multi-departmental team, but also to have access to round-the-clock services for preservation of tissue integrity^44^. Additional consideration includes legal requirements and ethics guidelines governing consent for autopsy procedures, and the lack of awareness of warm autopsy among academic oncologists or investigators^45^.

In this study, we preformed, to our knowledge, the first warm autopsy program in China. Through multidisciplinary teamwork, we collected eight metastatic tissues from a mCRPC patient throughout the warm autopsy within 2.5 hours after death. We comprehensively depicted the genetic and epigenetic cancer clonal evolution, and dissected the heterogeneous dynamics of the metastatic process of prostate cancer in this patient. Furthermore, we found that there was a hot spot in *CDKN1B*, which may be an important driver event to promote prostate cancer metastasis. The protein p27Kip1, encoded by *CDKN1B*, could inhibit tumor cell migration and invasion by binding to stathmin^46^. When *CDKN1B* was truncated, the loss of stathmin binding region (amino acids 170-198) and downregulation of expression may be the mechanism that promotes tumor metastasis^47^. In prostate cancer, the predominant variant types of *CDKN1B* were truncating mutation and deep deletion, which have been associated with increased risks of metastasis^48,49^. Finally, the metastatic phylogenetic tree we constructed further supported the previous viewpoint that genetics and epigenetics are concordant in tumor evolution^24,50,51^.

The Changhai Warm Autopsy Team (CWAT) was dedicated to collect samples from end-stage disease and identify resistant and metastatic mechanisms of cancer. With the support of CWAT, here, we not only performed warm autopsy, but also explored and established the internationally accepted standards in cancer field in China. This study and the warm autopsy program have important clinical and research implications. Our previous work in building the Chinese Prostate Cancer Genome and Epigenome Atlas revealed genomic and epigenomic signatures that were distinct from those of Western patients^15,33^. Investigating how such signatures evolve during tumorigenesis and metastasis can be critical to the understanding of disease mechanism and development of therapy. Warm autopsy provides an important platform to collect appropriate specimens for achieving the goal of reconstructing the steps of tumour evolution from initiation to metastasis^52^. Moving forward, we will initiate warm autopsy programs in large cohorts, conduct no fewer than 10 warm autopsies per year. Meanwhile, we are designing clinical trials to integrate longitudinal genomic and epigenomic studies to track cancer resistance and dissemination, following the TRACERx Consortium model^53,54^. Furthermore, we have initiated programs to raise awareness of warm autopsy among patients, their relatives, and the general public.

## Conclusions

We performed the first warm autopsy in China. We call on Chinese urologists and oncologists to promote warm autopsy of prostate cancer. We found the driver gene *CDKN1B* that is associated with the metastasis of prostate cancer, and the mechanism of this gene’s role in metastasis awaits further investigation. We also delineated the congruence of genetic and epigenetic clonal evolution during tumor metastasis. In conclusion, the warm autopsy process and standard and analysis pipelines we have established will pave the way for more investigators to apply them to the clinical diagnosis and treatment of prostate cancer.

## Data availability

The sequencing data in BAM format are available at the Genome Sequence Archive (GSA) for Human at the BIG Data Center (http://bigd.big.ac.cn/gsa-human/), Beijing Institute of Genomics with the accession number PRJCA010385.

## Acknowledgements

We sincerely thank the patient and his family. We wish to expressly thank Zhengmao Lu (Department of General Surgery), Changwei Yang (Department of Orthopaedic Surgery), Yuan Zou (Department of Neurosurgery), Chenguang Li (Department of Cardiothoracic Surgery) and Yeqing Lu (Operating Room) for the critical help implementing the warm autopsy. We would like to thank Guiling Ding (Department of Pathology) and Cong Wu (Biological Sample Bank) for coordinating the pathology review, tissue processing and labeling.

## Authors’ contributions

Conceptualization, XG and JL; Formal Analysis, WZ, TW and ZW; Investigation, BL, LC and XY; Writing—Original Draft, WZ and HS; Writing—Review & Editing, YW, MQ and XL; Funding Acquisition, XG and JL; Resources, YY, QY and FL; Supervision, ZL. All authors read and approved the final manuscript.

## Ethics approval and consent to participate

This study was approved by the ethical board at Changhai Hospital (No: TMEC2014-001, No: CHEC2019-165). Informed consent was obtained from all participants involved in the study, and all experiments were conducted in line with the principles of the Declaration of Helsinki.

## Consent for publication

There are no individual person’s data from all participants involved in the study.

