## Supplementary_Materials_and_Methods for "Genomic and epigenomic evolution of metastatic prostate cancer: the first warm autopsy in China"

**SUPPLEMENTAL INFORMATION**

1. Supplemental materials and methods

2. Supplementary figure legends

1. **Supplemental materials and methods**

**DNA and RNA extraction and quantification**

Genomic DNA was extracted using the DNeasy Tissue Kit (Qiagen) according to the manufacturer’s protocol. For formalin-fixed paraffin-embedded (FFPE) biopsy samples, DNA was extracted using the QIAamp DNA FFPE Tissue Kit (Qiagen). Qubit fluorometer and the Qubit dsDNA HS Assay Kit (Invitrogen) were used to access the DNA quality and yield. RNA was extracted using TRIzol reagent (Invitrogen) according to the manufacturer’s protocol. Qubit RNA Assay Kit and a Qubit 2.0 Fluorometer (Life Technologies) were used to measure the RNA concentration. RNA purity was measured using a NanoPhotometer spectrophotometer (IMPLEN, Germany).

**Library generation and sequencing**

A total of 800 ng of genomic DNA and 200 ng of RNA per sample were used as input material for library preparation. WGS, WGBS and RNA-seq library construction were performed as previously described^1^. NimbleGen SeqCap EZ Human Exome Library was used for exome capture before sequencing. WES libraries were generated as previously described^2^. Finally, WGS (with 188.67X average coverage on 12 samples), WES (with 1068.5X average coverage on 8 samples), WGBS (with 48.24X average coverage on 7 samples), and RNA-seq (with 24.55 Gb average bases data on 8 samples) libraries were sequenced on the Illumina HiSeq X TEN platform (2×150-bp paired-end reads). The following statistics were calculated in Additional file 1: Table S1: total number of reads; rate of sequencing error; percentage of reads with an average quality score greater than 20; percentage of reads with an average quality score greater than 30; and GC content of the sequences.

**RNA-seq** **processing**

The quality control check on raw reads was performed with FastQC v.0.11.3, and the clean reads were obtained by removing reads containing adapters and low-quality reads using Trimmomatic v0.33^3^. Clean reads were aligned to the reference human genome (build hg19) utilizing STAR v.2.7.6a^4^. StringTie v.2.1.4^5^ was used to assemble transcripts, and raw count data per gene was calculated using HTSeq^6^. Transcripts per million (TPM) were calculated for each gene using the GENCODE release 29^7^.

**ctDNA targeted sequencing and processing**

The ctDNA libraries were enriched with a pan-cancer ctDNA panel (Supplementary Table S4) provided by Shanghai Biotecan Medical Laboratory Co., Ltd. The ctDNA samples were subjected to preparation in the library according to the manufacturer’s protocol. Libraries were sequenced on the Illumina HiSeq X TEN platform (2×150-bp paired-end reads). Sequencing reads were aligned to the hg19 reference using BWA^8^, and GATK was used to adjust and refine the alignments. Somatic mutations were detected using Mutect2^9^. The candidate mutations were reviewed manually on the IGV.

**Sequencing data analysis for WGS and WES**

Raw paired-end reads in FastQ format were quality estimated by FastQC v.0.11.3 and pre-processed to high-quality using Trimmomatic v0.33^3^ to remove adapters and perform trimming. Sequencing reads were aligned to the reference human genome (build hg19) using BWA-MEM (Burrows-Wheeler Aligner) v.0.7.8^8^ (parameters -M -t 24). Unaligned reads that passed Illumina’s quality filter were retained. BAM files were processed and sorted from SAM files using Samtools v1.8, and Picard v1.130^10^ was used to combine data from multiple libraries and perform deduplication. Indel Realignment and Base Quality Score Recalibration in Genome Analysis Toolkit (GATK) v3.2 (https://www.broadinstitute.org/gatk/) were used to adjust and refine the alignments.

**WGBS processing**

FastQC v.0.11.3 was used to check the quality of the raw reads and perform basic statistics on the quality of the clean reads. Trimmomatic v0.33^3^ was used to pre-process raw reads and perform trimming. The clean reads were mapped to the reference human genome (build hg19) using Bismark (v.0.22.1)^11^ with the parameters (Bowtie2 –dovetail –score_min L,0,-0.2 –nucleotide_coverage). Duplicate reads that aligned to the same genomic region were removed using the deduplicatebismark command. The command of bismark_methylation_extractor in Bismark was used to extract cytosine methylation levels from the de-duplicated reads (–comprehensive –ignore_r2 18 –ignore 2 –bedGraph –no_overlap –report). The methylation and total read counts per CpG were calculated by the Coverage2cytosine command. All subsequent analyses were performed on CpGs with coverage of at least five. The bisulfite conversion rate of each sample was evaluated as the percentage of thymine sequenced at cytosine reference positions in the lambda DNA.

**Cell culture and cell transfection**

The Cancer Cell Line Encyclopedia (CCLE, <https://sites.broadinstitute.org/ccle/>) database was used to check the RNA expression of *CDKN1B* gene in prostate cancer cell lines, and 22RV1 cell line was selected for subsequent functional experiments validation. 22RV1 was maintained in RPMI medium 1640 (Gibco, USA) containing 10% fetal bovine serum (Gibco, USA) and 1% antibiotics (penicillin-streptomycin solution, Gibco, USA). The plasmids for transient transfection of short hairpin RNA (shRNA) for gene knockdown were purchased from Shanghai GenePharma (SuperSilencing™, catalog #C01001). We designed two shRNAs targeting *CDKN1B* (CDKN1B-sh1 and CDKN1B-sh2) and one negative control empty vector (NC-shRNA). The expression vector plasmid contained pGPU6/GFP/Neo, in which pGPU6 was an RNAi vector without screening marker, and could simultaneously express GFP protein to aid in the evaluation of transfection efficiency. Transfection of plasmids was performed by using GP-transfect-Mate (Shanghai GenePharma, catalog #G04009). After 48 hours of transfection, transfection efficiency was observed under a fluorescence microscope, and total RNA was extracted from the cells for real-time quantitative PCR (qRT-PCR).

The total RNA extraction from cells was performed using the MolPure® Cell/Tissue Total RNA Kit (Shanghai Yeasen, #19221ES50), and reverse transcription to cDNA was performed using the Hifair® III 1st Strand cDNA Synthesis SuperMix kit (Shanghai Yeasen, # 11141ES60), according to the manufacturer's instructions. qRT-PCR detection was carried out using a customized qRT-PCR quantitative assay kit (Shanghai GenePharma, # E23001) with dye-labeled primers and GAPDH as a internal reference gene.

**Cell proliferation and cell cycle analysis.**

The proliferation of 22RV1 cell line was evaluated using the CCK-8 assay kit (Beijing Solarbio, Cat. No. CA1210), following the manufacturer's instructions. Approximately 2,000 cells were added to each well of a 96-well plate to form a 100 μL system, and incubated for 24 hours prior to transfection. At 24, 48, and 72 hours post-transfection, the culture medium was removed from each well using a pipette, and 100 μL of CCK-8 detection reagent was added to each well. The plate was then incubated for 2 hours in a cell culture incubator, and the absorbance at 450 nm was measured using a microplate reader (Tecan, Switzerland). All experiments were performed in triplicate.

Cell cycle profiles was performed using the Cell Cycle and Apoptosis Detection Kit (Shanghai QiHaiFuTai, #C005-200), following the manufacturer's instructions. Cells were seeded in 6-well plates at a density of 2.5×10^5^ cells/mL, and after 24 hours of incubation, transfection was performed. After 72 hours of further incubation, the supernatant was discarded and the cells were digested. The propidium iodide (PI) staining solution was added to each sample, and the cells were gently resuspended by pipetting and incubated in the dark at 37°C for approximately 30 minutes. After staining, the cells were analyzed using a flow cytometer. All experiments were performed in triplicate.

**Scratch assay**

Cells were seeded at a density of 5×10^5^ cells/mL in each well of a 6-well plate. After incubating in a cell culture incubator for 24 hours, the fully grown cells in the 6-well plate were moved to a laminar flow hood. A sterilized ruler was placed above the plate as a reference, and a cross-shaped scratch was made in the plate by dragging the pipette tip along the ruler. The cells were washed twice with PBS to remove the scratched cells. Next, 1mL serum-free medium was added, and the distance of the scratch was recorded under a microscope at 0 hours. The cells were then transfected routinely, and photographs were taken 72 hours after culturing. Three replicates were performed.

**Migration and invasion assay**

For the cell migration and invasion assay, the cell suspension was added to serum-free medium to achieve a concentration of 2.5×10^5^ cells/mL after cell digestion. 700 μL of culture medium with 10% fetal bovine serum was added to each well of a 24-well plate, and then Transwell chambers (Corning, USA) were inserted. For invasion experiments, Transwell were pre-treated with Matrigel matrix. 200 μL of the prepared cell suspension was added to each Transwell, and then incubated for 6 hours in a cell incubator until the cells completely adhered to the wall. The plate was then removed from the incubator, and the transfection reagent mixture was added to each Transwell, followed by continued incubation in the cell incubator for 72 hours.

The Transwell was gently removed with tweezers, and then washed and gently wiped to remove the upper layer of cells. Cells were fixed with 4% paraformaldehyde and stained with crystal violet in a 24-well plate. After washing several times with PBS, the Transwell chamber was allowed to dry completely, and then photographed and counted under an inverted microscope. Three replicates were performed.

**2. Supplementary figure legends**

**Supplementary Fig. 1 Dynamic change of *BRAF* and *TP53* mutation in ctDNA.**

After applying Vemurafenib, the allele fraction of the *BRAF* mutation was significantly dropped (left), and the clonal mutation of *TP53* also fluctuated (right).

**Supplementary Fig. 2 The pathology of metastatic sites procured by warm autopsy.**

The pathology (100x magnification) of specimens used for sequencing (A) and those used only for pathological review (B).

**Supplementary Fig. 3 Fraction of each mutational signature in the genome.**

Stacked bar charts indicate the known mutation signature as defined by Alexandrov et al (https://cancer.sanger.ac.uk/signatures/).

**Supplementary Fig. 4 Sanger sequencing validation of the truncating mutation of *CDKN1B*.**

The figure shows the sequencing chromatogram of the the Sanger sequencing validation in bone sample (LV2M) and metastases sample (PVLN).

**Supplementary Fig. 5 Subclonal composition and clonal evolution of the metastases.**

(A) Anatomic distribution and tissue type of study samples. (B) The figure shows the composition profiles of the mean cancer cell fraction of mutations in each cluster inferred by PyClone. (C) The firgure represents the clonal dynamics over time of the individual tumor, which corresponds to Fig 4C and D. The sphere of cells shows the clonal admixture or subclonal population in each sample.

**Supplementary Fig. 6 CDKN1B mutations in public data.**

(A) We roughly counted *CDKN1B* mutations harbored in all prostate cancer patients (n=9510) in cbioprotal (https://www.cbioportal.org/). The alterations of *CDKN1B* gene was founded in approximately 4% of prostate cancer samples, and the truncating mutation and deep deletion were the predominant variant types. The legend is exactly the same as cbioprotal. (B) *CDKN1B* expression level in normal tissues, primary tumors and metastases (harbored truncating mutation) in the RNA-seq data. The public RNA-seq data of 136 primary tumor/normal sample pairs were from the Chinese Prostate Cancer Genome and Epigenome Atlas (CPGEA), and no tumors harbored *CDKN1B* truncating mutation. Non-biological batch effects were inspected using Principal Component Analysis (PCA). The TPM, transcripts per million. *P* values were determined by two-sided Mann–Whitney U-test (***, *P* value < 0.001). (C) Validation of *CDKN1B* expression using RNA (top, N=333) datasets in TCGA. The data are publicly available from the cbioportal website (https://www.cbioportal.org/). The figure indicates that the expression of truncated *CDKN1B* is significantly downregulated compared to the unmutated samples. The y-axis represents log2(value+1) transformed RNA seq expression data. *P* values were determined by two-sided Mann–Whitney U-test (***, *P* value < 0.001; **, *P* value < 0.01; *, *P* value < 0.05; ns, *P* value > 0.05).

**Supplementary Fig. 7 The effects of downregulating *CDKN1B* expression on 22RV1 cells.**

(A) *CDKN1B* expression in prostate cancer cell lines from the CCLE database (N=9), with the y-axis representing the log2(value+1) transformed TPM values. (B) Cell cycle profiles tested by flow cytometry in 22RV1 cells. (C) CCK-8 assay to measure the viability of 22RV1 cells after *CDKN1B* knockdown. (D) Scratch assay to evaluate the migration ability of cells. P values were determined by two-sided Student’s T-test.

**Supplementary Fig. 8 DNA methylation level and epigenetic aberrations across different metastatic tumors.**

(A) Distribution of CpG methylation levels in seven tumors and three normal samples. (B) The dendrogram represents the similarity of their methylation profiles based on the clustering of the samples. The used distance method and clustering method are "correlation" and "ward" respectively. (C) The firgure, related to Fig 5F, shows the difference in methylation levels of CpG island in promoter region of known prostate cancer driver genes. Only numbers that the difference between each tumor and three normal prostate samples more than 15% are shown in the heatmap. (D) Epigenomic clonal evolution tree that added three normal prostate specimens (N1, N2, and N3). Lengths of trunks and branches were inferred using the top 1% of CpG sites with the greatest difference between different tumor regions. The method and color coding are the same as Fig 6C.
