## Supplementary_figures for "Genomic and epigenomic evolution of metastatic prostate cancer: the first warm autopsy in China"

Supplementary Figure 1

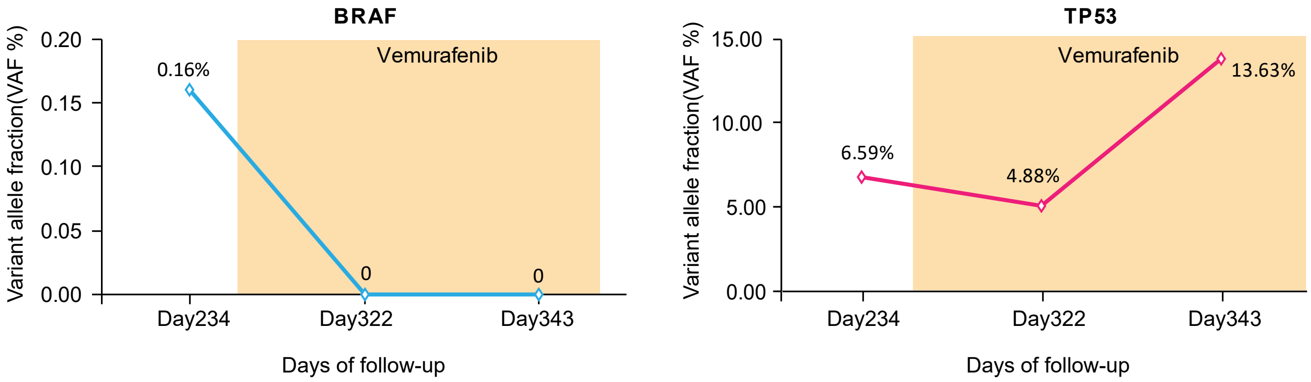


**Supplementary Fig. 1 Dynamic change of *BRAF* and *TP53* mutation in ctDNA.**
After applying Vemurafenib, the allele fraction of the *BRAF* mutation was significantly dropped (left), and the clonal mutation of *TP53* also fluctuated (right).

Supplementary Figure 2

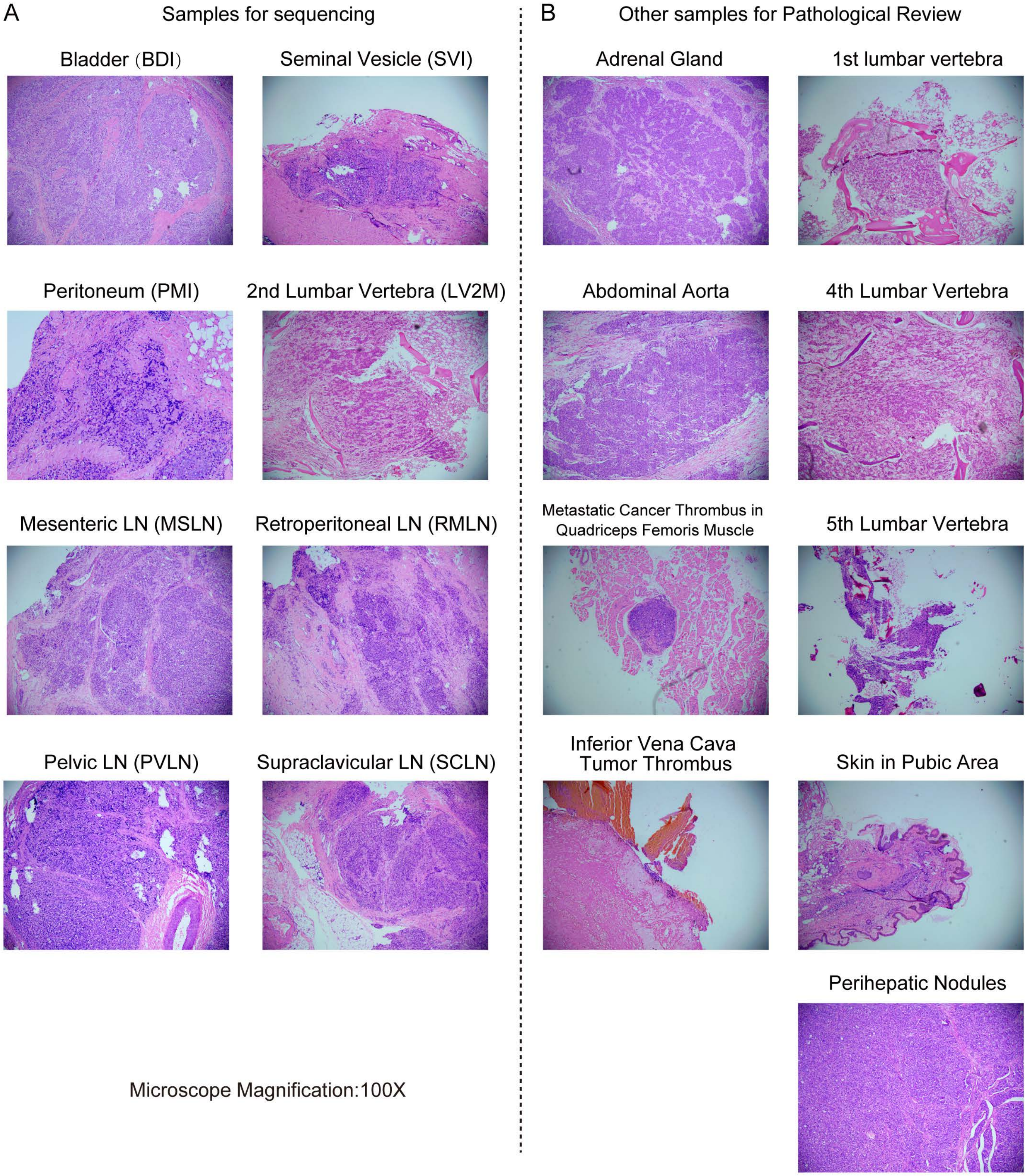


**Supplementary Fig. 2 The pathology of metastatic sites procured by warm autopsy.**
The pathology (100x magnification) of specimens used for sequencing (A) and those used only for pathological review (B).

Supplementary Figure 3

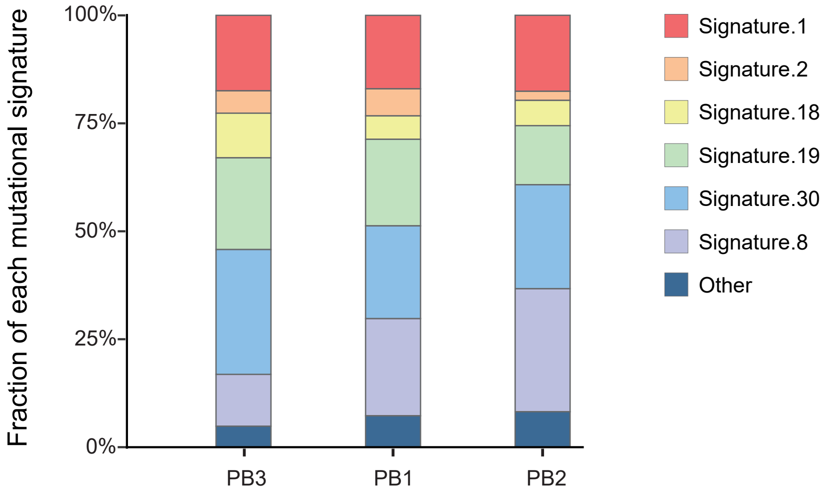


**Supplementary Fig. 3 Fraction of each mutational signature in the genome.**
Stacked bar charts indicate the known mutation signature as defined by Alexandrov et al (https://cancer.sanger.ac.uk/signatures/).

Supplementary Figure 4

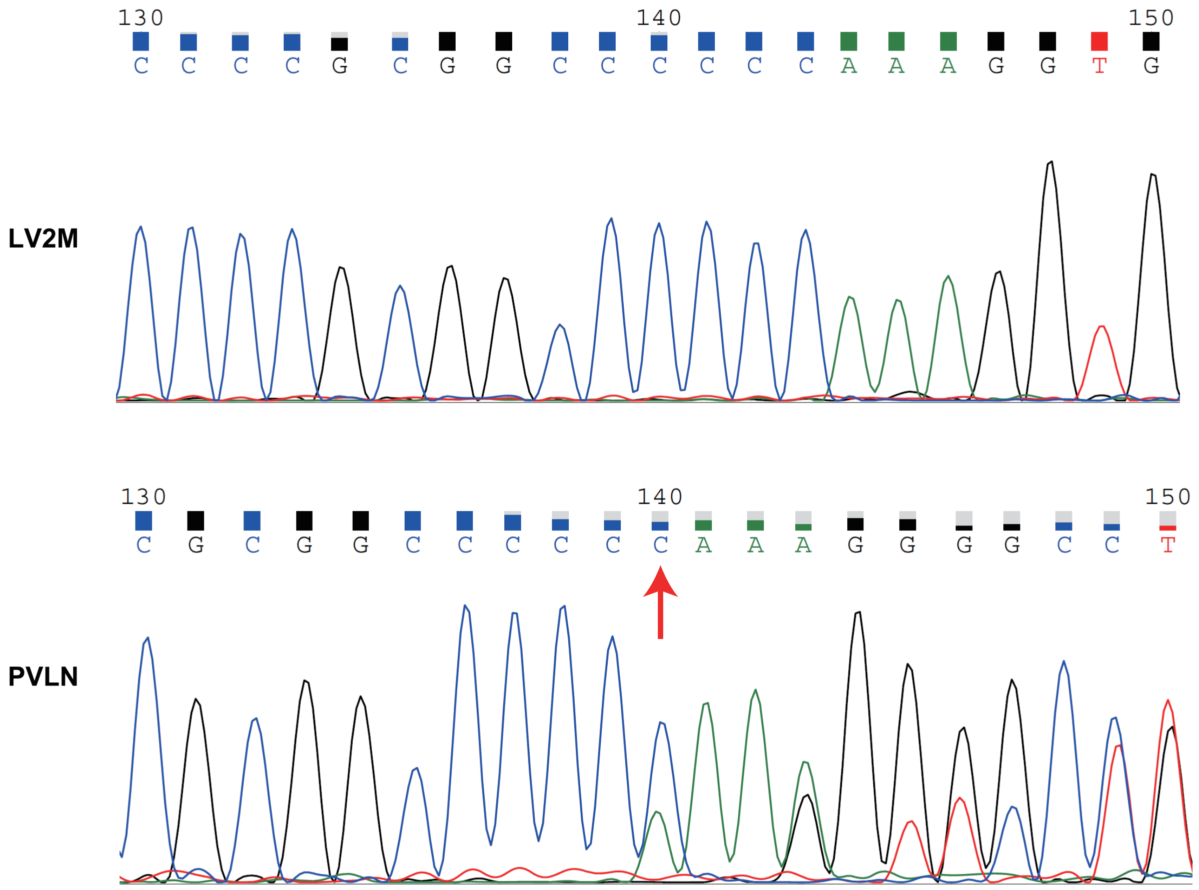


CDKN1B p.(Gly97Valfs*22) : chr12_12871053_12871053_C_-

**Supplementary Fig. 4 Sanger sequencing validation of the truncating mutation of *CDKN1B*.**
The figure shows the sequencing chromatogram of the the Sanger sequencing validation in bone sample (LV2M) and metastases sample (PVLN).

Supplementary Figure 5

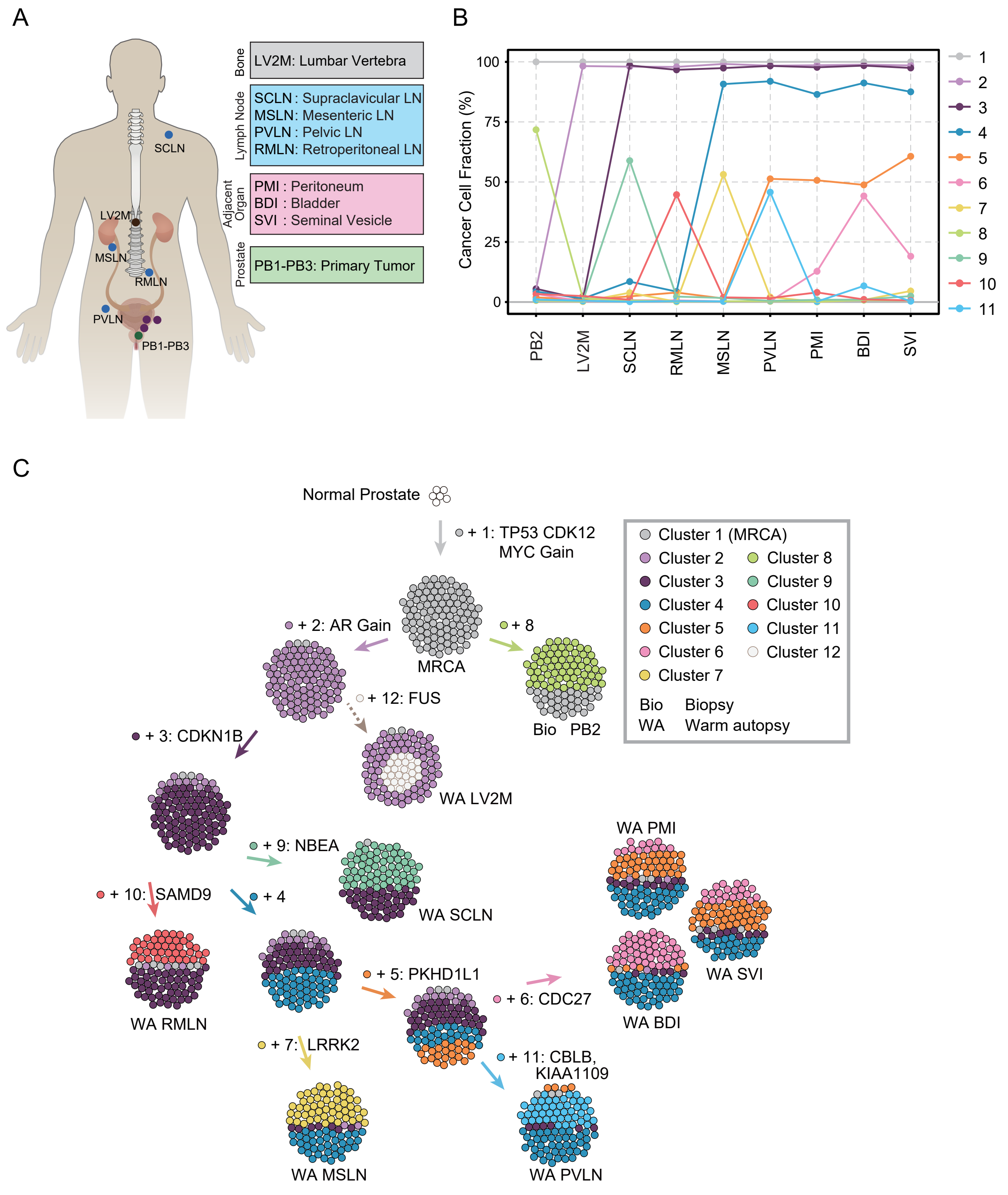


**Supplementary Fig. 5 Subclonal composition and clonal evolution of the metastases.**
(A) Anatomic distribution and tissue type of study samples. (B) The figure shows the composition profiles of the mean cancer cell fraction of mutations in each cluster inferred by PyClone. (C) The firgure represents the clonal dynamics over time of the individual tumor, which corresponds to Fig 4C and D. The sphere of cells shows the clonal admixture or subclonal population in each sample.

Supplementary Figure 6

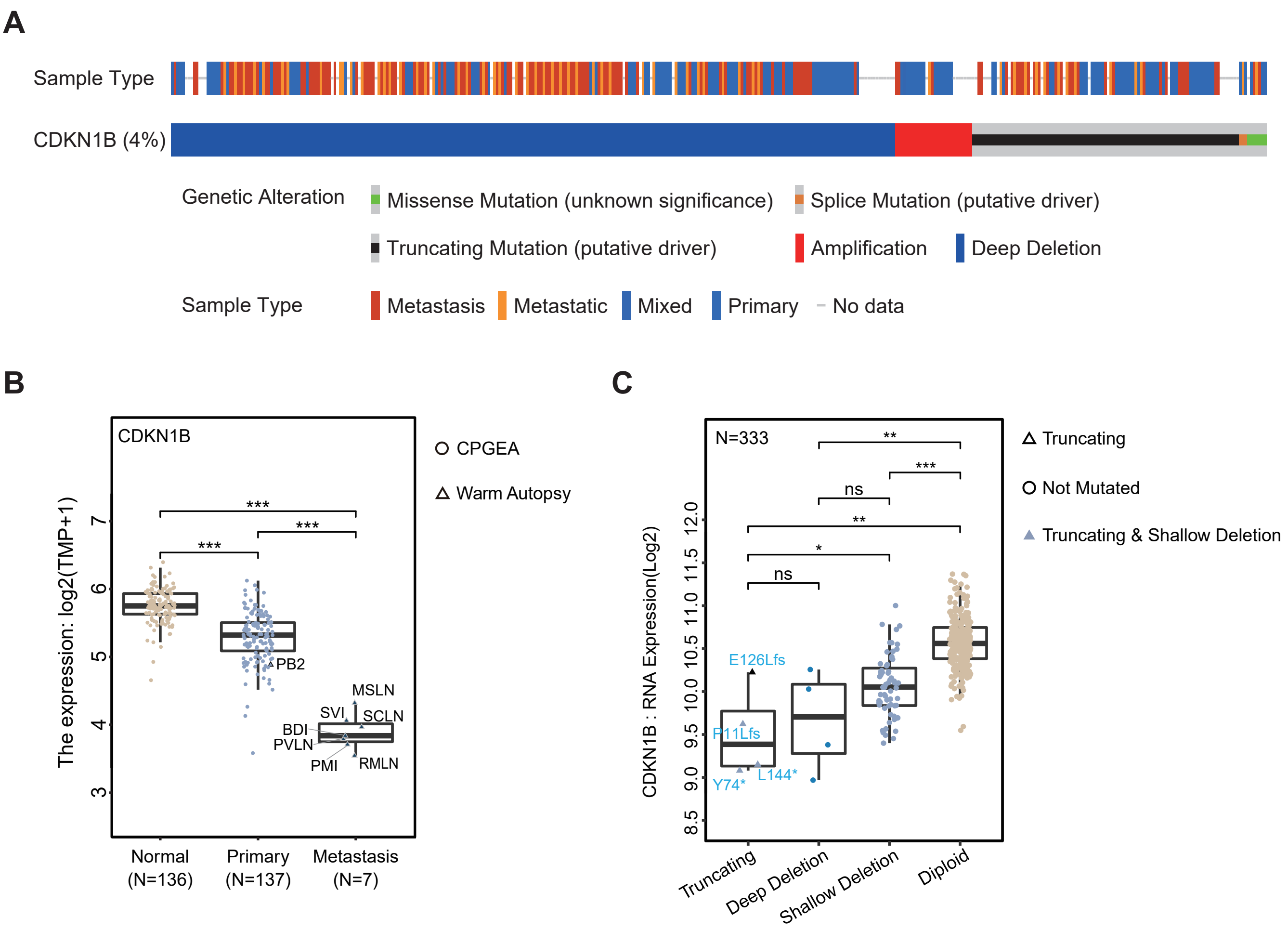


**Supplementary Fig. 6 *CDKN1B* mutations in public data.**
(A) We roughly counted *CDKN1B* mutations harbored in all prostate cancer patients (n=9510) in cbioprotal (https://www.cbioportal.org/). The alterations of *CDKN1B* gene was founded in approximately 4% of prostate cancer samples, and the truncating mutation and deep deletion were the predominant variant types. The legend is exactly the same as cbioprotal. (B) *CDKN1B* expression level in normal tissues, primary tumors and metastases (harbored truncating mutation) in the RNA-seq data. The public RNA-seq data of 136 primary tumor/normal sample pairs were from the Chinese Prostate Cancer Genome and Epigenome Atlas (CPGEA), and no tumors harbored *CDKN1B* truncating mutation. Non-biological batch effects were inspected using Principal Component Analysis (PCA). The TPM, transcripts per million. *P* values were determined by two-sided Mann–Whitney U-test (***, *P* value < 0.001). (C) Validation of CDKN1B expression using RNA (top, N=333) datasets in TCGA. The data are publicly available from the cbioportal website (https://www.cbioportal.org/). The figure indicates that the expression of truncated *CDKN1B* is significantly downregulated compared to the unmutated samples. The y-axis represents log2(value+1) transformed RNA seq expression data. *P* values were determined by two-sided Mann–Whitney U-test (***, *P* value < 0.001; **, *P* value < 0.01; *, *P* value < 0.05; ns, *P* value > 0.05).

Supplementary Figure 7

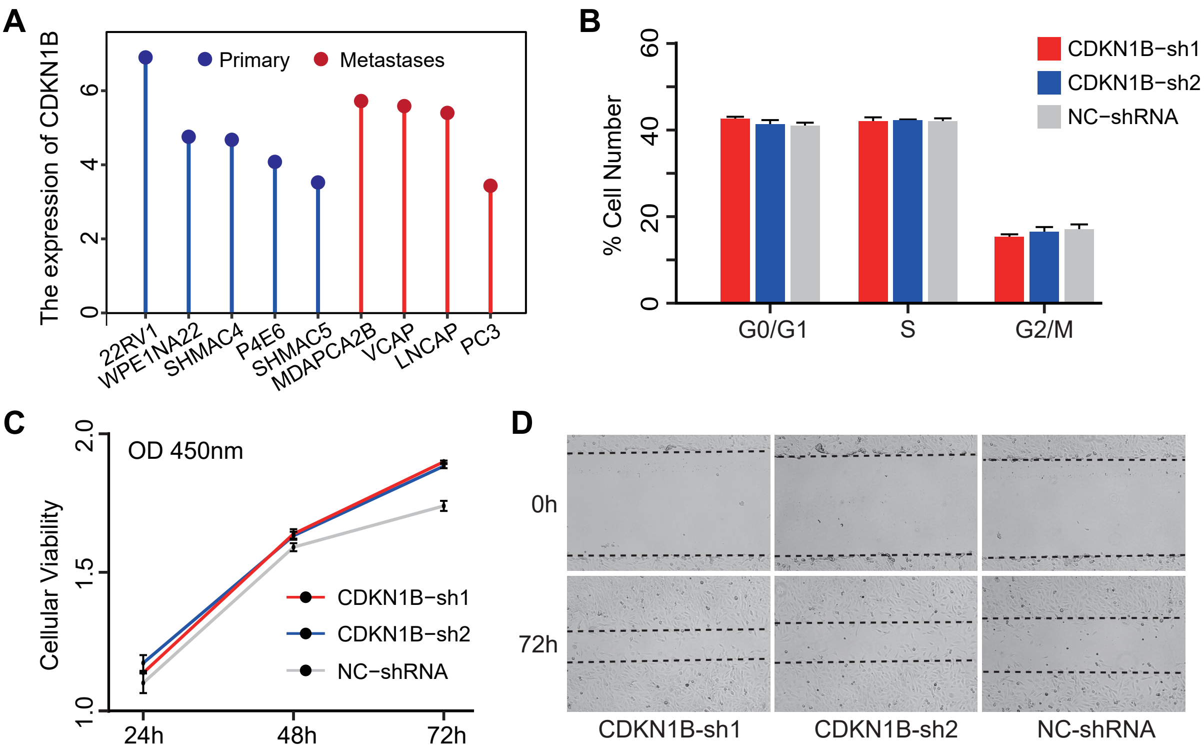


**Supplementary Fig. 7 The effects of downregulating *CDKN1B* expression on 22RV1 cells.**
(A) *CDKN1B* expression in prostate cancer cell lines from the CCLE database (N=9), with the y-axis representing the log2(value+1) transformed TPM values. (B) Cell cycle profiles tested by flow cytometry in 22RV1 cells. (C) CCK-8 assay to measure the viability of 22RV1 cells after *CDKN1B* knockdown. (D) Scratch assay to evaluate the migration ability of cells. P values were determined by two-sided Student's T-test.

Supplementary Figure 8

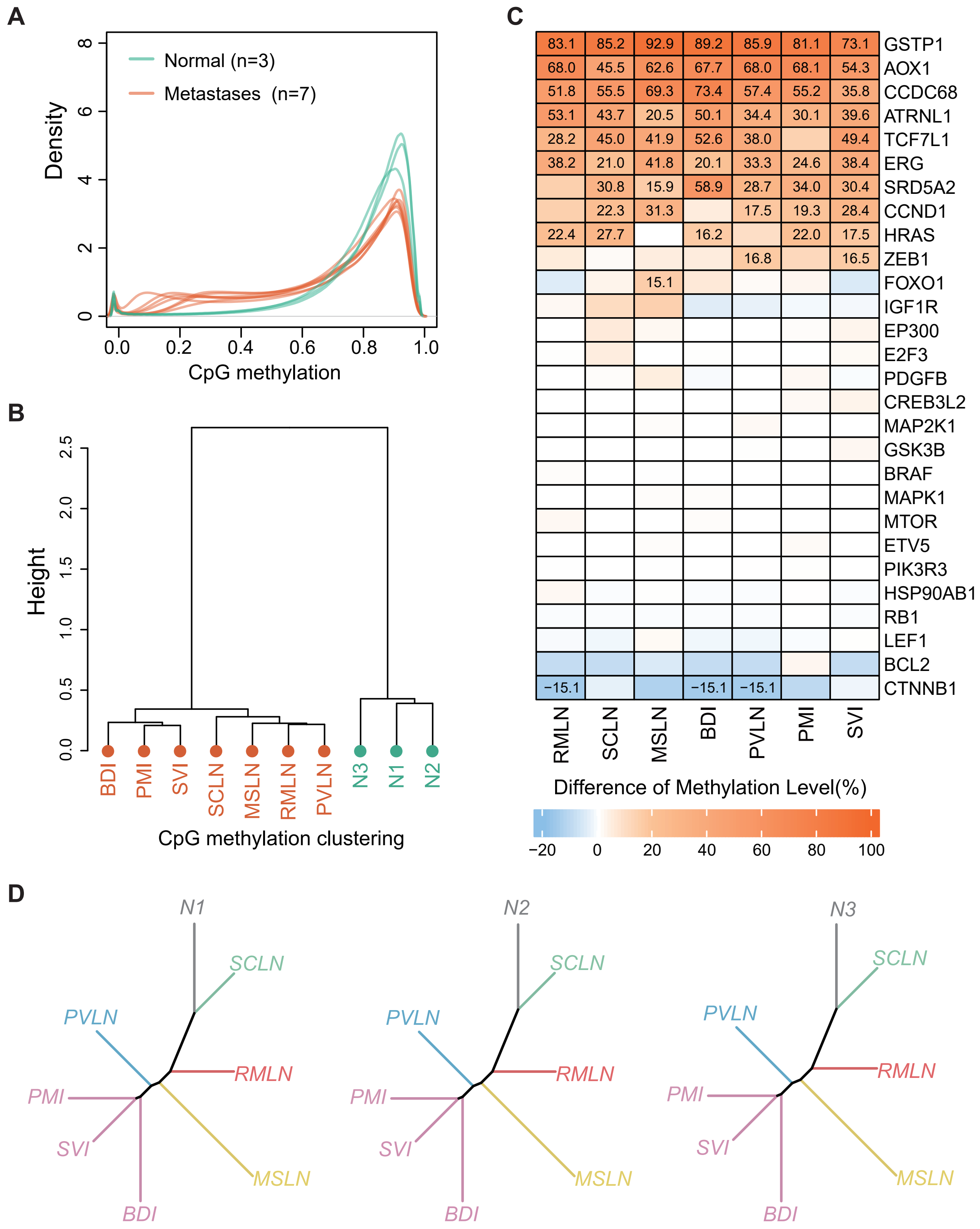


**Supplementary Fig. 8 DNA methylation level and epigenetic aberrations across different metastatic tumors.** (A) Distribution of CpG methylation levels in seven tumors and three normal samples. (B) The dendrogram represents the similarity of their methylation profiles based on the clustering of the samples. The used distance method and clustering method are "correlation" and "ward" respectively. (C) The firgure, related to Fig 5F, shows the difference in methylation levels of CpG island in promoter region of known prostate cancer driver genes. Only numbers that the difference between each tumor and three normal prostate samples more than 15% are shown in the heatmap. (D) Epigenomic clonal evolution tree that added three normal prostate specimens (N1, N2, and N3). Lengths of trunks and branches were inferred using the top 1% of CpG sites with the greatest difference between different tumor regions. The method and color coding are the same as Fig 6C.
